# Public attitudes to COVID-19 vaccines: A qualitative study

**DOI:** 10.1101/2021.05.17.21257092

**Authors:** Simon N Williams, Kimberly Dienes

## Abstract

**OBJECTIVE:** To explore public attitudes to COVID-19 vaccines in the UK, focused on intentions and decisions around taking vaccines, views on ‘vaccine passports’, and experiences and perspectives on post-vaccination behavior.

**DESIGN:** Qualitative study consisting of 6 online focus groups conducted between 15^th^ March – 22^nd^ April 2021.

**SETTING:** Online video conferencing

**PARTICIPANTS:** 29 adult UK-based participants

**RESULTS:** Three main groups regarding participants’ decision or intention to receive a COVID-19 vaccine were identified: (1) Accepters, (2) Delayers and (3) Refusers. Two reasons for vaccine delay were identified: delay due to a perceived need more information and delay until vaccine was “required” in the future. Three main facilitators (Vaccination as a social norm; Vaccination as a necessity; Trust in science) and six barriers (Preference for “natural immunity”; Concerns over possible side effects; Distrust in government; Perceived lack of information; Conspiracy theories; “Covid echo chambers”) to vaccine uptake were identified. For some delayers, vaccine passports were perceived to be a reason why they would get vaccinated in the future. However, vaccine passports were controversial, and were framed in three main ways: as “a necessary evil”; as “Orwellian”; and as a “human rights problem”. Participants generally felt that receiving a vaccine was not changing the extent to which people were adhering to COVID-19 measures.

**CONCLUSIONS:** Overall, positive sentiment toward vaccines was high. However, there remains a number of potential barriers which might be leading to vaccine delay in some. ‘Vaccine delay’ might be a more useful and precise construct than vaccine hesitancy in explaining why some may initially ignore or be uncertain about vaccination invitations. Vaccine passports may increase or ‘nudge’ uptake in some delayers but remain controversial. Earlier concerns that vaccination might reduce adherence to social distancing measures are not borne out in our data, with most people reporting ongoing adherence and caution.

## Background

One year after the novel coronavirus (COVID-19) pandemic led to its first lockdown in March 2020, the UK vaccination programme is continuing at pace. Vaccine uptake has been a key focus throughout the pandemic, in both policy and public discussions (UK Government 2021a; The Guardian 2021). Vaccination is widely considered as a critical tool to bring the pandemic under control (World Health Organization). Debates around vaccination are multi-faceted and complex. In this paper, we focus on public views on three major issues related to COVID-19 vaccination: (1) vaccine uptake, specifically facilitators and barriers to uptake (including reasons for vaccine hesitancy); (2) vaccine passports; (3) behaviour and adherence to COVID-19 measures (non-pharmaceutical interventions) following receipt of a vaccine (one or both doses).

In the UK, as of 29 April 2021, approximately 34 million people had received a first dose, 14 million of which had also received a second dose (UK Government 2021b) This equates to approximately 59-70% of the adult population across the four countries. Globally, vaccine uptake intentions appear mixed, although a recent systematic review of 13 countries suggests that overall a majority (60%) intend to take a COVID-19 vaccine (Robinson et al 2021). However, latest data from the UK suggests that uptake promises to be considerably higher, with approximately 90% of adults reporting they have either received a vaccine or would be likely to have one if offered (ONS 2021a; Ipsos Mori 2021a). However, despite the overall high uptake of, or intention to receive, COVID-19 vaccines, there are a number of disparities in terms of who is more or less likely to receive, or be willing to receive, a vaccine. Office of National Statistics (ONS) (2021b) data shows that vaccine uptake differs by ethnicity, religion or socio-economic status, with for example, lower vaccination rates amongst Black and Asian Minority Ethnic (BAME) individuals. In terms of intentions, the systematic review by Robinson et al found that being female, younger, of lower income or education level and belonging to an ethnic minority group were consistently associated with being less willing to be vaccinated (Robinson et al 2021).

One reason for the variation in vaccination rates across socio-demographic groups is variation in vaccine hesitancy. Vaccine hesitancy can be defined as “the delay in acceptance or refusal of vaccination despite availability of vaccination services” (MacDonald et al 2015). As has been acknowledged, vaccine acceptability exists on a spectrum and it is important to distinguish vaccine hesitancy - where individuals may have limited information or who may have genuine concerns about the safety or efficacy of a specific vaccine – from anti-vaccination (anti-vax) sentiment – where individuals are wholly opposed to vaccination per se (Burgess et al 2021). In this paper, we draw on The World Health Organization’s SAGE Working Group on Vaccine Hesitancy’s ‘continuum of vaccine hesitancy’ model, which sees vaccine views to be set on a continuum between full acceptance of vaccines with no doubts, through to complete refusal with no doubts (MacDonald et al 2015). Vaccine hesitancy is seen as a heterogenous group in-between, including those who “delay” acceptance (i.e. do not get it when first offered or according to schedule). Additionally, determinants of compliance with vaccination have been understood in terms of the “3 C’s model”, of complacency (i.e. low perception of risk from the disease), convenience (i.e. practical or logistical barriers to access vaccination) and confidence (i.e. lack of trust in the safety or efficacy of the vaccine) (SAGE 2014).

Research suggests that younger adults, female adults, BAME individuals, adults with lower formal educational qualifications, and those living in the most deprived areas are all more likely to report vaccine hesitancy (than young males, White British individual, or adults living in the least deprived areas respectively) (ONS 2021b; Robertson et al 2021). Recent survey evidence suggests that the most common reasons for vaccine hesitancy include: worries over side effects, worries over long term effects on health, as well as concerns over its efficacy (ONS 2021a).

Qualitative research on public views on COVID-19 vaccines is limited, with, at time of writing, little-to-no qualitative research focused on UK public vaccine views. One study in Canada on overall attitudes to public health measures to reduce COVID-19 transmission found that many participants felt that vaccines were a means to “get back to normal life” while some were hesitant due to a lack of confidence in the potential efficacy of the vaccine and concerns over side-effects (Bentham et al 2021). One study focused on India found a range of mixed views across people’s knowledge, attitudes, perceptions and concerns (Kumari et al 2021). Further qualitative research can play a vital role in understanding COVID-19 public views (Teti, Schatz & Liebenberg 2021) since it can provide more nuanced perspectives and an in-depth understanding of the reasons behind these perspectives.

As the vaccination program is rolled out across the UK, but as disparities - both in terms of domestic and international vaccine coverage – exist, the topic of “vaccine passports” has become increasingly debated. Essentially, vaccine passports can be considered a specific type of “immunity certificate” - through which people are able to prove they are significantly less likely to carry and transmit COVID-19 to others, by virtue of the fact they have tested negative or in this case, received a vaccination. In the UK, as globally, the issue of vaccine passports has been controversial subject of debate in both policy and the media, with those opposed arguing that passports would for example infringe upon civil liberties, and those in favour arguing for example that it might enable people to feel more “confident” in public venues and could “unlock the economy” (Walker 2021; Drury 2021). They have also been the subject of academic debate, including a number of opinion pieces which debate their potential use, including issues of access, equity and ethics (Osama, Razai & Majeed 2021; Hall & Studdert 2021). At present, comparatively little empirical academic research has been done on public attitudes to vaccine passports. Although survey research is beginning to explore the issue (Ipsos Mori 2021b), there is a dearth of qualitative research on the subject.

Another question that has been debated in relation to vaccines is whether or not receiving a vaccine has impacted people’s adherence to COVID-19 social distancing measures (non-pharmaceutical interventions). A recent review drew on evidence from other diseases as well as small number of surveys related to COVID-19, and found that some of those who have been vaccinated will show reduced adherence to personal protective behaviours (SPI-B 2021). However, it also concluded that there is limited evidence relating to possible changes of behaviour due to vaccine rollout (SPI-B 2021). At time of writing, little-to-no qualitative evidence on people’s experiences and perspectives on post-vaccine behaviour and adherence exists.

In this paper, we explore public views on COVID-19 vaccines in the UK, focusing on three main questions: (1) What are the major barriers and facilitators for vaccine uptake? (2) What are people’s views on vaccines passports and how do they frame these views? (3) Has having a vaccine (one or two doses) changed the extent to which people are adhering to COVID-19 measures (non-pharmaceutical interventions)?

## Participants and Methods

Data from this study came from the COVID Public Views Study – a collaboration between researchers at Swansea University, the University of Manchester and the World Health Organization. The COVID-19 Public Views Study is an ongoing longitudinal mixed-methods study using panel focus groups and surveys during the pandemic (commenced March 2020). Further details, including methodological information, about the study can be found in previous publications (Williams et al 2020a; Williams et al 2020b). Participants are invited to take part at frequent intervals during the pandemic, usually to coincide with important developments or milestones during the pandemic. In this paper, we report on data from six online focus groups with 29 participants from the COVID Public Views Study, conducted between 15 March and 22^nd^ April 2021. Participants responded to a rapid invitation to take part in these focus groups in order primarily to discuss the issue of vaccines.

Participants were initially recruited to the study from March-July 2020, with a total of 53 participants enrolled into the study. Participants were all UK-based adults aged 18 years or older. Recruitment for the study took place via a combination of social media advertising (Facebook ads), snowball sampling (e.g. via Twitter, Facebook Groups), other online advertising (e.g. online ‘free-ads’ such as Gumtree) and other recruitment methods (e.g. via University press releases). Purposive sampling was used to seek as diverse a range of ages, genders, race/ethnicities, UK locations, and social backgrounds as possible. However, despite these attempts to recruit as diverse a sample as population, certain groups, in particular individuals from older age groups (over 50 years of age) were underrepresented in the final sample. However, in the context of vaccination uptake, the over-representation of younger age groups (18-49) can also advantageous, given at time of writing (April 2021) vaccination roll-out is starting to reach younger adults, where, as discussed above, it is hypothesized that uptake might be lower than amongst older age groups (50+). The limitations of the sample are discussed previously (Williams et al 2020a), and below, alongside some of the strengths of the method and sample. Full demographic summary details are provided in Table 1.

**Table 1:** Demographic details reported by participants.

| **Characteristic** | **N (%)** |
| --- | --- |
| *Gender* |  |
| Female | 11 (38) |
| Male | 18 (62) |
| *Age range* |  |
| 20s | 9 (31) |
| 30s | 8 (28) |
| 40s | 9 (31) |
| 50+ | 3 (10) |
| *Ethnicity* |  |
| White | 20 (69) |
| BAME | 9 (31) |

Online focus groups were necessary due to COVID-19 social distancing regulation, but have been seen to have benefits in general, as a means of eliciting public views from diverse and geographically dispersed participants (Tates et al 2009; Williams 2010). Each focus group (of 4-6 participants) met virtually via a web videoconferencing platform (Zoom) for approximately one hour. All focus groups discussed in the present paper were moderated by SW (a social scientist). The topic guide for the focus groups was initially developed using existing literature on vaccination public attitudes and vaccine hesitancy discussed above, as well as rapidly emerging research on COVID-19 public attitudes.

Given the fact that, as noted above, a key focus of both policy and public discussions at the time was the issue of vaccination (including facilitators and barriers to uptake, vaccine passports, and post-vaccine behaviour), it was also a key topic of discussion in the focus groups.

### Analysis

Data were analysed in accordance with a framework analysis approach (Pope 2000). Data were coded and organized according to themes and subthemes, using a data matrix (framework). Both authors (SW and KD) analysed the data. Coding was performed using NVivo (version 11.4.3, QRS). Both authors met regularly to discuss and consult with each other during the course of the analysis. The framework approach entails a number of processes. Firstly, authors familiarised themselves with focus group data by reading and re-reading transcripts. Authors then developed an analytical framework for the analysis, by coding a sample of transcripts. After agreeing on a provisional coding framework, remaining transcripts were coded (with additional themes being added as they emerged). Framework analysis can be primarily deductive or inductive, depending on the particular research question (Gale et al 2013).

The WHO SAGE Working Group on Vaccine Hesitancy’s ‘continuum of vaccine hesitancy’ (MacDonald et al 2015) was particularly important in shaping the basic framework for our analysis of the facilitators and barriers related to vaccine uptake, although we sought not to use it prescriptively and to allow new themes to emerge which did not fit this model. Due to the novel nature of the COVID-19 pandemic and COVID-19 vaccination, data analysis on the issue of vaccine passports and post-vaccine behaviour proceeded largely inductively, looking for emergent, novel themes within the data. The themes presented below represent our most prominent themes and are exemplified with sample quotations.

Ethical approval for the study was granted by Swansea University’s School of Management and College of Health and Human Sciences.

## Results

### Attitudes to vaccination

Amongst our participants, and building on existing literature on vaccine hesitancy (MacDonald et al 2015; Burgess et al 2021), we found three distinct groups of vaccine attitudes: (1) *Accepters* (2) *Refusers* (3) *Delayers*.

#### Accepters

One group, (n=15 (52%)), accepters, held largely positive views around COVID-19 vaccines, and who had either accepted a vaccine, or who were intending on accepting a vaccine when it was their time to be offered one. In line with MacDonald et al’s (2015) vaccine hesitancy continuum, accepters fell into two sub-types: those who expressed little to no reservation about their decision to accept a vaccine (“full accepters”) and those who had accepted, or were intending to accept, a vaccine, but who also expressed concerns or reservations about their decision (“accept but unsure”).

Full accepters tended to frame their decision as something that was quite “normal” to them (who had “had all my vaccines” - see *vaccination as a social norm* below):

> “I had my first vaccine at the end of February, and I had no problems getting the vaccines or any concerns about the efficacy of the vaccines … It was something I need to do to protect my family and loved ones … and the people I engage with on a da-to-day basis in the community … It was an easy decision to make, I’ve had all my vaccines all my life, because I see the value.” (Participant 44, Female 30s)

Full accepters also framed vaccination as a collective act, and their decision as something that would “protect” others as well as themselves (especially vulnerable others) (“I don’t want to hurt anyone else” (Participant 19, Male, 20s). This included younger accepters and those who saw themselves as not being at risk personally:

> “I had my first one about a month ago … I was practically queuing up as soon as I heard … My biggest thing throughout the whole pandemic to protect others, not necessarily myself, because I’ve never seen myself as being particularly at risk but now I am statistically less likely to catch and pass it on” (Participant 6, Male, 20s)

However, while some accepters (n=8 (28%)) (like Participant 6, above), were confident in their decision, others (n = 7 (24%)) were more likely to have concerns in spite of ultimately deciding to get vaccinated (akin to the Vaccine Hesitancy Continuum’s ‘accept but unsure’ vaccine attitude group) (MacDonald et al 2015):

> “I was a bit suspicious. I suppose it’s only natural, everyone’s a little worried because you think it’s only been done in a year, but the science today compared to years ago is outstanding isn’t it, so they have more methods of testing to see if its effective, so I wasn’t really worried, and I’m glad I have got it” (Participant 42, Female 30s)

Accepters, including those in the “accept but unsure” category, were more likely to emphasise facilitators, including a *trust in science* and *vaccination as a necessity* (see below).

#### Refusers

A second group (n=2 (7%)), refusers, held largely negative views around COVID-19 vaccines. They quite clearly stated that they had decided that they were not going to get a vaccine. Unlike the delayers (below) they did not give an indication that they might change their mind for “the foreseeable future”:

> “I’ve not had the vaccine and, to be honest with you, I don’t have any intention of getting the vaccine like in the foreseeable future, I just don’t see the point”. (Participant 3, Male, 20s)

Refusers tended to frame vaccination as an individual act rather than a collective act; as a personal choice (“It’s a choice I’ve made” (Participant 8, Female, 40s)).

Refusers were less likely to frame their vaccination as something that would protect others and saw it as something that was primarily useful for those most vulnerable from serious COVID-19-related illness, for example older age groups (see *preference for natural immunity* below). Refusers emphasised the three main barriers, including a *preference for natural immunity concerns over side effects* and *distrust in government* (see below).

#### Delayers

A third group (n=12 (41%)), hesitants, were unsure of whether or not to get the vaccine. Corresponding with Burgess et al (2021), these participants can be distinguished from those unequivocally and ideologically opposed to vaccination – that is *refusers*. Unlike the refusers, they had not yet made – that is, they were *delaying* (MacDonald et al 2015) - their decision as to whether they would have a vaccine. Some delayers had already turned down or ignored their vaccination appointment invitations. However, unlike refusers they characterized their current position as temporarily and potentially subject to change – that is they did not want the vaccine *yet* but were aware that they *might* in future:

> “I wouldn’t take it straight away… I’d rather just wait and make my own decision at my own pace really” (Participant 15, Male, 20s)
>
> “I hear people saying like yeah I’m definitely having it, but then when you talk to people more, they start saying well I want more people to have it before me” (Participant 28, Female, 20s).

Some delayers tended to emphasise a need for more information about the efficacy and safety of the vaccine as a main reason for their delay (see *perceived lack of information* below):

> “I’m still holding out. I’ve been sent several letters and loads of text messages telling me that I needed to go for the vaccination, but I would rather wait at least four months to see exactly where it is all going… I don’t want to be sort of pressured into something that I might regret” (Participant 15, Male, 40s)

These participants tended to be concerned about the speed at which vaccines had been developed and were more likely to be concerned over the possible longer-term side effects (see *concerns over possible side effects* below):

> “I am a little bit nervous because I do feel like it’s all been rushed through incredibly quickly … have they done all the tests? I wanted to ensure that a lot more people had it before it was my turn, because I kind of figured that if there’s a problem, we’ll know” (Participant 50, Female, 50+)

Despite the feeling of “pressure”, these participants tended to perceive themselves as having agency in their choice of whether to get vaccinated.

Other refusers however, tended to focus on a second common reason for delay was that participants felt they would ultimately “have” to get the vaccine and so were waiting until they needed to do so:

> “Personally, I don’t think I want to get it until I’m actually forced to do it” (Participant 7, Female 20s)
>
> “I don’t intend on taking it, unless it’s a must, unless, I heard you have to go on holidays, but right now I am going to refuse it, until the very last moment. But I know at some point I will have to take it” (Participant 18, Female, 30s)

These participants tended to be quite reluctant, or even opposed, to getting a COVID-19 vaccine, but believed that they would have to get one in order to be able to undertake certain activities in future, particularly travelling internationally. Here, vaccination tended to be viewed as an individual act (“I have the say and right what I should accept into my body and nobody else should have a say” (Participant 22, Female, 20s)). They tended to perceive themselves as having little or no agency in regard to whether they would ultimately have a vaccine (see *vaccine passports* below). The act of delaying may therefore be construed as a way to re-capture the sense of control or agency over their lives that many felt they had lost during the pandemic (Williams et al 2020c).

### Facilitators to uptake (reasons for getting vaccinated)

#### Vaccination as a social norm

Social norms are a means through which health behavior and decisions can be influenced during the pandemic (Bavel et al 2020). Accepters tended to discuss COVID-19 vaccination in relation to a personal willingness to be vaccinated in the past and in general (“I’ve had vaccines all my life for what is needed” (Participant 12, Male, 40s)), as well as in relation to a culture (the UK) within which vaccines were normative (including the fact that vaccinations were a “part of normal life” from a young age):

> “Especially in this country we are used to being vaccinated for various things, in childhood and then, especially because I used to travel a lot in my work, and depending on the country I was visiting, there were a few vaccines that you need to get” (Participant 11, Male, 50+)
>
> “For me, I’ve never been one to say no to a vaccine. Its part and parcel - as a child you are given various vaccines, and you don’t necessarily have a choice, but then when you have your boosters, you just go ahead and do it and to me that’s part of normal life” (Participant 10, Male, 40s)

For some, vaccination for COVID-19 was becoming more normative over the course of the pandemic. This, they felt, was influencing some to move from being hesitant or even opposed to get vaccinated towards being vaccinated:

> “Initially when it [vaccine] came out a lot of the [front line-workers] on my team were like ‘nope, not going to get it’ but that changed one-by-one, they started to go for it, and now I think there is only a handful who haven’t gone for it … I don’t know why that changes, but initially they had reservations … as I said its being monitored by senior management but they have got to ask have you had it or not, they are not asking why you haven’t had it, but I don’t know if that is something at some point, you know you have to justify your grounds as to why you haven’t” (Participant 5, Female, 30s)

For this participant, in their workplace, there was an implied expectation that they would, in the future, need to get the vaccine or justify why they hadn’t. This informal scrutiny of vaccine uptake may be functioning as an example of ‘norm-nudging’ (Biccheri & Dimant 2019). In this participant’s workplace, although vaccination was not obligatory or coercive, and the reasons for ‘opting-out’ were not required, the implication was that opting-in was the default or normative position. The expectation justification for opting out would be necessary in the future may have led to the change in decision (i.e. individuals opting to get vaccinated now, despite not initially wanting to, because they felt they would either be required to in the future or would have to justify their (non-normative) decision not to).

#### Vaccination as a necessity

Vaccination as a social norm closely related to a second facilitator – vaccination a necessity. Participants discussed three main ways in which vaccination would be necessary or inevitable. These were framed either positively or negatively (related to whether participants were accepting of vaccines or not).

Firstly, accepters in particular spoke of vaccination as being the “the way for the to “get back to normal” (Participant 20, Male, 20s) or “the only way forward, to get on with our lives” (Participant 12, Male, 40s). Secondly, a number of participants, including accepters and hesitants, described an “acceptance” of the fact that they would need regular vaccinations or ‘boosters’ as a result of the continued emergence of new variants of the virus:

> “My parents and I have already accepted that we will require boosters, they will be a fact of life, probably for the next four or five years, and it will be the same as our flu jab that we get every year” (Participant 2, Male, 40s)

In particular, concerns over new and emerging variants were cited as a reason as to why vaccination was not perceived to be a “one-off”, and so “regular” vaccination was seen to be necessary with “no other way out”.

> “The virus is going to keep mutating, there is going to be different variants, I think that’s why the government is so hot on this vaccine because it is not going to stop. … It’s something that we’re going to have to live with this is not going to die out anytime soon … there is no other way out … we are going to have to go for our top-ups with the vaccine” (Participant 5, Female, 30s)

It is worth noting however, that refusers were more likely to see the possibility of regular vaccination as a deterrent to getting vaccinated currently:

> “There’s a concern that, if one takes the vaccine have a variant of a new variant emerges. That could be resistant to the vaccine and then there’s a question of do you have to keep taking vaccines to protect yourself against each single various oh I don’t really have any intention of getting the vaccine” (Participant 3, Male, 20s)

Thirdly, immunity certificates or “vaccine passports” were also cited by some participants as a potential facilitator to vaccine uptake. Discussion of vaccine passports elicited a range of views, discussed in more detail below (see *vaccine passports*). However, one frequent theme, particularly amongst those who were currently hesitant was that vaccine passports were seen as a reason as to why they felt they would ultimately “have to” get a vaccine:

> “I’m two minds really about whether to get the vaccine or not but I have a feeling people will be pressurized into having it through the vaccine passports or certificates, whether that could be for travelling or getting jobs, so I think people will be indirectly forced into getting it” (Participant 7, Male, 40s)

#### Trust in vaccine science

Accepters were more likely to exhibit a higher trust in science, and to frame science as being relatively independent from government:

> “I chose to be vaccinated because, not that I trust the government, but I trust the medicine and the science behind it it’s not the government that produces the vaccine. The government just pays for it, and if the government is prepared to spend money on something that might prevent me from getting seriously ill, I’m quite happy to take that**”** (Participant 11, Male, 50+)

Accepters were also more likely to associate the vaccination program with the health service (NHS):

> “I know that the vaccine was developed really quickly … but I have faith in the health system and its testing” (Participant 10, Male, 40s)

Although some accepters expressed some concern at the speed at which the vaccines had been developed, they tended to contextualise this in terms of the fact that science was now more advanced than with many previous vaccines:

> “I was a bit suspicious; I suppose it’s only natural, everyone’s a little worried because you think it’s only been don’t in a year, but the science today compared to years ago is outstanding isn’t it, so they have more methods of testing to see if its effective, so I wasn’t really worried, but I’m glad I have got it” (Participant 25, Female, 30s)

They also contextualised the speed of vaccine development in terms of what they perceived as rigorous testing, and the fact that there had been considerable scientific, medical and financial focus and investment in the vaccine development. This for them meant any potential risks from unforeseen issues due to safety or efficacy were likely to be minimal and outweighed by the benefits of the vaccination program:

> “I believe it’s been so rigorously tested and it’s the only thing spoken about medically for the last 12 months and so the risk for me was just so minimal and benefits outweighed any risk for me” (Participant 6, Male, 20s)

### Barriers to uptake (reasons for vaccine hesitancy)

#### Preference for ‘natural immunity’

Some participants argued that one of the reasons they were either hesitant about or did not want to get vaccinated was because they preferred to “fight” the virus “naturally”:

> “For me personally I am not sure I would go for the vaccine… I just hope I have a strong immune system so I can fight the virus. We have this in-built immune system within our bodies …give them a chance to operate” (Participant 7, Male 40s)

In discussing their decision, refusers tended to frame COVID-19 as a disease which tended not to affect young and “healthy” people:

> “I don’t have any intention of getting the vaccine like in the foreseeable future, I just don’t see the point, because the virus is mainly fatal like to those who are like middle aged. (Participant 3, Male, 20s)

In doing so, they also emphasised their own healthiness as a reason as to why they didn’t need the vaccine, and drew comparisons to the fact they hadn’t needed vaccines in the past for other diseases:

> “It just does not make sense to me to take a vaccine, it’s like a flu vaccine. Ive never ever taken a flu vaccine, because I don’t get the flu.” (Participant 8, Female, 40s)

As noted above, refusers framed vaccination as an individual act rather than a collective act, and argued that the lack of personal benefit was outweighed by the perceived risks posed from potential side effects:

> “I have no intentions of taking it, and I have focused a lot on my health over the years, I’m the healthiest I’ve ever been and I just don’t see I don’t the reason for me to take it … because from what I’ve read, there are risks with it so.” (Participant 8, Female, 40s)

#### Concerns over possible side effects

One of the main reasons for vaccine hesitancy was a concern over side effects – something that accounted for why some people were delaying the decision to get vaccinated:

> “I probably will have it [a vaccine], but I want to wait to see if people turn into zombies first. I’ll wait until a few hundred thousand have had it first” (Participant 28, Female, 20s).

Although the above quote was tongue-in-cheek, it was indicative of a wider concern over potentially unforeseen, longer-term, side effects. Hesitants tended to frame these concerns in relation to what they knew about how vaccines were “normally” developed. They discussed how comparatively quickly COVID-19 vaccines had been developed and emphasized how not enough time had passed to be able to know long-term side effects:

> “I do have like some concerns about how quickly they developed this vaccine, because most vaccines take like you know six five to six years to test and to make sure that you know they’ve seen all the side effects. But with this vaccine I still have that reservation that maybe it’s been too quick, and they’ve not really teased out all the long-term effects.” (Participant 3, Male, 20s)

Other hesitants focused on short-term side effects or risks. In particular, recent reports of potential blood clots linked to vaccines were cited by a number of vaccine-hesitants. Interestingly, these participants were aware that any causal link between blood clots and vaccines had not been clearly demonstrated, or that any potential risk between vaccines and blood clots was considerably small. Nevertheless, they continued to cite blood clots as a cause for concern and framed their hesitancy in relation to it as an example of potential side effects (including unknown side effects that may emerge in the future):

> “Right now, I am going to refuse it, until the very last moment. I feel like I’m a guinea pig. I don’t know if you heard the news that they have stopped one of the vaccines because there has [sic] been cases of blood clots of something. I know it’s a very, very, very, very tiny percentage but I feel like if I wait till the very last more [information] can come out”. (Participant 9, Female, 30s)

Some participants discriminated between vaccines, with those who did expressing personal concern, or observed concern in others, over the AstraZeneca vaccine, which had been the focus of blood clot controversies in the media. For example, one participant, who was vaccine hesitant, stated that although they accepted the Pfizer vaccine, they would doubtfully have accepted the AstraZeneca vaccine:

> “Now I’ve had it I feel okay about it, but I think that’s because with the Pfizer it doesn’t seem to be any negative reports, whereas with AstraZeneca there seems to be a lot of mixed communications, and I don’t think there is a lot of fault with the vaccine, it’s just I don’t think the company is very good at kind of being truthful and that makes people a bit doubtful … I don’t think I would have done it [had the AstraZeneca vaccine] (Participant 24, Female, 50+).

Another participant described how they had heard of others specifically opting out of vaccination, once they discovered they were to get the AstraZeneca vaccine:

> “My aunt had hers and she said there was a huge queue in the surgery … but every single person that was offered the one that begins with an A [AstraZeneca] were actively declining it and walking out and she witnessed in ten minutes about 15 people turning up, being told what they were getting and walking out” (Participant 21, Female, 20s)

On the other hand, accepters, including those who had taken the vaccine with reservations (cf. McDonald et al.’s (2015) ‘accept but unsure’ group), were much less likely to be concerned over both short- and long-term side effects. They framed the issue in terms of, a cost-benefit analysis, where the, perceived low risks were outweighed by the high societal benefits. They also made comparisons to what they saw as other equally rare side effects of common medications:

> “I know there is this whole issue around blood clots, but people really need to get a grip, because you know, people die on a yearly basis from taking paracetamol, plenty of women die from blood clots as well …from taking the pill … that so the benefits outweigh the risks most definitely” (Participant 12, Male, 40s)
>
> “I’m quite worried about not any side effects now, but like maybe in ten years’ time … but then it seems like the risk from the vaccine is less than the risk of say getting long Covid … so the vaccine is the lesser of two evils” (Participant 24, Female, 50+)

#### Distrust in government

Refusers and some hesitants tended to have less confidence in vaccine science, less trust in government, and were more likely to frame COVID-19 vaccine science as being closely linked to, or even compromised by, political or economic interests. Some justified their distrust in relation to historical controversies or examples of clinical iatrogenesis:

> “I think a lot of my concerns are because of the government because I just don’t trust them at all (Participant 29, Female, 50+)
>
> “The government doesn’t have a very good track record with the sense that there was the Thalidomide tablets that were given to pregnant women back in the 60s, the blood transfusions that were imported from people volunteers in state penitentiaries in America that were contaminated, brought into the United Kingdom there’s been several vaccinations given to toddlers … that came from America I think about 1000 children have died…. I’m not one to trust governments, they tend to rush into things.” (Participant 15, Male, 40s)

Although hesitants generally recognised that the vaccines had been tested, they remained concerned, or “skeptical” that testing had been done as extensively or for as long as was necessary. They tended to see vaccination as something that was still being tested (in the community):

> “Even though there has been a lot of testing done, I still feel skeptical and quite scared to get it … technically it’s still in the testing phases even though it’s been approved, and so until I’m actually forced to do it, I don’t think I want to” (Participant 7, Female, 20s).

#### Perceived lack of information

A perceived lack of information was a major factor for why some participants were either refusing or delaying vaccination:

> “Whatever is going on with the vaccine, I don’t know, it really is a minefield of information” (Participant 8, Female, 40s)

For many the questions were technical in nature, some of which were legitimate scientific questions stemming from the fact that COVID-19 was still such a new disease. For example, uncertainty over post-infection immune response and efficacy against new variants were some of the reasons for why participants were reluctant or unsure of whether a vaccine would “work” and thus was worth having or not:

> “There’s a concern that, if one takes the vaccine and a variant of a new variant emerges that could be resistant to the vaccine then there’s a question of do you have to keep taking vaccines to protect yourself against each single variant? I don’t really have any intention of getting the vaccine.” (Participant 3, Male, 20s)
>
> “I am not against the vaccine, [but] for me there are so many unknowns, because It is so new there are so many questions that I want to ask, like if you have had Covid do you still need, in terms of the antibodies you have, or do you still need the protection from the vaccine? I’m very much in the middle, so once I get those answers, I will be leaning more towards getting it” (Participant 5, Female, 30s)

Some accepters also felt that they were unsure about their decision because of a lack of information being provided to them:

> “There wasn’t a lot of information … I mean I never really understood how they make them and stuff and it was never explained to me and so it’s just like I was going to get something put in my arm, I was like ‘oh God!’” (Participant 19, Male, 20s)

#### Conspiracy theories

A number of participants referred to conspiracy theories and misinformation, with refusers and delayers more likely to discuss them uncritically (that is they did not acknowledge them as conspiracy theories or misinformation as such. Conspiracy theories were also discussed in relation to the previous theme of a distrust in government:

> “I mean distrust in government … the things that don’t seem to add up. I mean we have got the pharmaceutical companies, several of them creating a vaccine some kind of race … and it’s just a win-win for them, if just everyone gets a vaccine and people can’t think for themselves … it a big agenda” (Participant 7, Male, 40s)

Amongst the Black and Asian Minority Ethnic participants in this study, most were critical of the circulation of conspiracy theories. However, many of them did also discuss how they had heard or seen conspiracy theories and misinformation in their communities (“It’s weird how it afflicts the Black community in terms of like the social media and WhatsApp conspiracy theories in circulation” (Participant 2, Male, 40s). Some related the lower uptake in vaccination to the existence of ‘folk wisdom’ about what might help promote health or even protect against COVID-19:

> “A few months back India was number two in the number of cases and deaths from Covid and one fine day it just vanished. So, everyone is trying to ask what is being done differently in India … and I guess that mindset is being transferred to racial communities here [in the UK]. So people are discussing our [Indian] food habits, and we do eat a lot of spicy food and spices so I’ve actually seen people talking about saying that ‘ok it’s our food which is different … *there are people even saying avoid the vaccines, stick to your spices, your curries and you’ll be fine*, so that is worrying but that is a topic which is being widely discussed in our Asian community” (Participant 4, Male, 30s)
>
> “My father sent me something about someone sent him about lung capacity. If you hold your breath for X amount of time, then a you’re protected from Covid and you don’t need to worry about the vaccine, really bizarre stuff” (Participant 2, Male, 40s)

However, a number of BAME participants who linked the lower uptake in their communities to a distrust in government (see above) and thus a distrust in the vaccines. a distrust in vaccine science. One example was the rumour that vaccines were being “tested” for side effects first amongst BAME patients:

> “I think just from my experience is that there’s a lot of conspiracies that I’ve heard about. Because I mean I identify myself as a Black British person and so within my Community yeah that I’ve heard a lot, of just not trusting the vaccine … somebody sent me a video about [UK Government Health Secretary] Matt Hancock …suggesting that the vaccine was tested first amongst the BAME group.” (Participant 5, Female)

These participants also discussed the issue of distrust as a wider issue, accounting for why there “is some cynicism in these communities” (Participant 2, Male, 40s). This lack of trust was seen to stem from a lack of information (cf. *lack of information* above), which in turn was seen to be the result of a lack of engagement with BAME communities, as well as a perceived lack of government accountability for the disparities that BAME communities have experienced during the pandemic:

> “I just think within our community there needs to be a lot more education, especially if there are a lot of unknown questions that haven’t been answered – I don’t know if there are any forums where questions can be asked to medical professionals… questions that are not being asked in the media – that needs to be advertised more as to where we can go to ask those questions to be more equipped with the knowledge around these vaccines, rather than listen to these conspiracies which a lot of them is fake news…. A lot of it [lack of trust] stems from the government; a lot of how it [pandemic] has been handled is embarrassing, a high number of deaths were from the Black and Asian community and so that mistrust in government along with them not really putting their hands up just makes us even more anxious” (Participant 5, Female)

#### Covid ‘echo chambers’

One potential barrier to uptake is what might be seen as the emergence of ‘Covid echo-chambers’. Echo chambers can be defined as “polarized communities populated by like-minded” others and can be found particularly in online settings (Bessi 2016):

> “I also like interact quite a lot on the Internet and a lot of people that I speak to on the Internet, they say the same thing that you know they just have this concern about the vaccine.” (Participant 3, Male, 20s)

The existence of polarized views, on an emotionally charged subject led some participants to argue that ““everyone is extreme in their reactions” (Participant 26, Female, 20s):

> “It really divides those who do have the vaccine and those who don’t, and I see quite a lot on social media people who post like very critical things of people who are on the opposite side” (Participant 15, Male, 20s)

As a result, some hesitants felt ‘as though they didn’t ‘fit’ into either of the polarized attitude groups on vaccines, which may have been contributing to their uncertainty or hesitancy around whether or not to get vaccinated:

> “If you want to have the vaccine, good on you, I think you are braver than I am. But also, those people who don’t want to have the vaccine, I totally understand where you’re coming from. But there seems to be there are not many people with that attitude. Its either the people who [say] ‘don’t have the vaccine, it’s got 5G in it and the government are going to follow you and nobody should have it’ or you’ve got the people going ‘I’ve had the vaccine, and everyone should have the vaccine and you are stuffing it up for the rest of us” (Participant 26, Male, 30s)

Echo chambers are discrete and are characterised by a lack of communication across them. In our study we found evidence of a lack of communication between individuals with differing views on vaccines, for example between accepters and refusers (including so-called “Covid deniers”). For example, one participant, an accepter, described how having conversations with a family member about vaccines was difficult given the latter’s opposing views on Covid:

> “I’m happy to get the vaccine …but one family member isn’t keen on the vaccine, because they are just not convinced of the coronavirus in the first place… they think it’s not too big of a deal actually. I’ve not had a discussion with them about the vaccine because of his views on coronavirus in general” (Participant 6, Male, 20s)

Similarly, another participant, a refuser, described how she was reluctant to discuss the subject of vaccination with others, particularly those who she knew, or thought, may be in have strongly ‘pro-vaccination’ views:

> “I don’t really bring it up now in conversation now with anyone … A couple of people have asked me have I got it and I just said no I don’t want to get into discussions about belief because I am just taking each day as it comes. … I do find there is a lot more of two extreme sides…. I do find that most of the people I have spoken to who have gotten it are very passionate about it and are, not judgmental, but from their perspective they are afraid and so they can’t understand why others haven’t taken it (Participant 8, Female, 40s).

Some participants - refusers and some hesitants - were critical of the extent of advertising around vaccination. Some were skeptical that there needed to be so much, and implied that Covid was not as serious as had been made out by some. Refusers and hesitant were more likely to not have had COVID-19 themselves, nor know of many or any close others that had had it:

> “I have noticed there is so much advertising, and I wonder why does it need so much advertising? Why aren’t people dropping dead? … I don’t see evidence in our everyday life, I see it affecting people’s anxiety and people with cancer not getting treatment, but I don’t know anyone who’s died of Covid directly”” (Participant 8, Female, 40s)

One participant likened vaccination advertising to “propaganda”:

> “I feel like it is just propaganda all the time these days, and the vaccine roll out I just thought that was another farce” (Participant 24, Female, 50+)

From this perspective, what was seen as excessive advertising or propaganda may have been having a counter-productive effect, encouraging participants to disengage from the messages or to distrust them. Similarly, some BAME participants discussed how targeted advertising in their opinion were counterproductive because they “amplified” differences, thereby reifying the echo chambers:

> “For me we’re all British citizens. …and I think sometimes when you amplify these differences too much they can become a little counterproductive and I think it insulates and isolates people into their own new cocoons and I do worry about that I heard something about ‘Oh, we need a, we need a black doctor sort of you know, next to Chris witty to endorse this to the nation’ and I thought that was really bizarre” (Participant 2, Male, 40s)

### Vaccine passports

On the subject of vaccine passports, participants had mixed views on their value and acceptability. As noted above, a number of hesitants felt that vaccine passports would nudge or force them into ultimately needing a vaccine. Additionally, analysis revealed three main framings of vaccine passport: *vaccine passports as a ‘necessary evil’; vaccine passports as ‘Orwellian’; vaccine passports as a ‘human rights problem’*.

#### Vaccine passports as a ‘Necessary evil’

As noted, some participants (in the accepters group) felt that vaccine passports, particularly for international travel, were inevitable and would, they felt, ultimately lead them to get a vaccination:

> “It’s a good thing, you need to keep the people safe for holidays and thigs like that, to keep the people in other countries safe” (Participant 9, Female, 30s)

However, they did not necessarily see it as a positive policy. If fact, very few participants spoke unequivocally positively about vaccine passports. One framing that did emerge was the characterization of vaccine passports as a “necessary evil” – that is as undesirable but beneficial for public health:

> “I’m in hospitality, and how are restaurants, bars, clubs gonna open up again with any sort of safety any confidence without utilizing the fact that a certain portion of our society is going to be vaccinated and it’s that’s going to be a lot safer for those individuals in those establishments. So, to me I think it’s just it’s almost a necessary evil and it’s the way we’re going to go … I think it’s also going to save a lot of businesses and that will save you know; the economy and we all need the economy to be boisterous we all need a prominent economy”. (Participant 2, Male, 40s)

Specifically, it was argued that vaccine passports were a necessary way of “saving” the economy, as well as providing a sense of “safety” or “reassurance” (Participant 7, Male, 40s) for travellers and customers where they were required.

#### Vaccine passports as ‘Orwellian’

A second framing was of vaccine passports as “Orwellian”. Some participants (including accepters, hesitants and refusers) expressed concern over the issue of privacy and the fact that vaccine passports infringed on people’s privacy; they likened them to a dystopian government surveillance society (cf. George Orwell’s novel *1984*):

> “[I’m] a hundred percent against vaccine passports, I personally would rather just have the PCR tests. … it is somewhat Orwellian … I’m very, very concerned about things like surveillance and privacy” (Participant 3, Male, 20s)

Perhaps linking to previous themes – distrust in government and conspiracy theories - some participants were sceptical that “there’s another agenda there somewhere” (Participant 8, Female, 40s).

#### Vaccine passports as a “human rights problem”

Some participants, across all three groups, were strongly opposed to vaccine passports as a matter of ethics; some argued, simply that they were “wrong” (Participant 8, Female, 40s). The matter was framed in terms of “human rights”, with some arguing that vaccine passports would infringe on people’s human rights by taking away people’s freedom of choice, particularly for those individuals in society that weren’t able to get the vaccine (for example, on health grounds).

> “A lot of people don’t want to get vaccinated, and for them it will be a problem, a human rights problem” (Participant 40, Male, 40s)
>
> “For the UK, will that be imposing on our human rights, especially to go to places like the supermarket or the cinema or things like that? I feel under immense pressure in a way. It’s not deprivation of liberty, but it’s sort of saying that if you don’t follow the government’s guidelines, if you don’t have a certificate, you won’t be accepted. It’s about not having a freedom of choice relating to whether you want to take the vaccine or not, we are just continually pressured to take the vaccine and everything will be ok … because a lot of people won’t be able to consent to the vaccine for whatever reason.” (Participant 13, Male, 40s)

## Discussion

Our findings explore qualitatively the reasons and meanings behind three main issues: people’s decisions or intentions around getting vaccinated; people’s views on vaccine passports; people’s behaviour after receiving a vaccine. In keeping with the ‘continuum of vaccine hesitancy’ model (MacDonald et al 2015) we found three main groups of participants, based on their decision or intention to receive a COVID-19 vaccine: *Accepters*; *Delayers;* and *Refusers*. In order to explain these different attitudes, we identified three facilitators (*Vaccination as a social norm; Vaccination as a necessity; Trust in science*) and six barriers (*Preference for “natural immunity”; Concerns over possible side effects*; *Distrust in government; Perceived lack of information; Conspiracy theories; Covid echo chambers*) to vaccine uptake. As with existing research (ONS 2021a; Ipsos Mori 2021a) outright opposition to, including refusal of, a COVID-19 vaccine was very low (7%) in our study, while positive sentiment towards vaccines - including those who have already accepted or plan to accept a vaccine – was very high amongst our participants (52%). However, vaccine hesitancy may still be an issue for many who have either not yet been offered a vaccine or who have not responded to their vaccination invitation (41%). We suggest that the concept of ‘vaccine delay’ is a more precise and more useful means through which to understand why some people are reluctant or unsure as to whether or when they will receive a COVID-19 vaccine. We found two main reasons for the existence of vaccine delay. Firstly, some were delaying due to a perceived need for more information. Secondly, others were delaying until they were “required” to be vaccinated.

Our findings can be understood in relation to broader conceptual models of vaccine hesitancy, including the World Health Organization’s ‘Three C’s’ model (SAGE 2014) and the SAGE Working Group on Vaccine Hesitancy Determinants Matrix (MacDonald et al 2015) specifically to the case of COVID-19. Our findings suggest that, as per the ‘Three C’s’ model, particularly Confidence, and Complacency were major factors explaining vaccine hesitancy (Convenience was less relevant in our study, in part due to the rapid rollout being relatively consistently and successfully rolled out across the UK via the National Health Service (Baraniuk 2021)). For example, ‘confidence’ in the efficacy and safety of the vaccines was a major facilitator or barrier depending on a person’s perspective, especially in relation to the extent to which they trusted science or government (and the extent to which they saw the latter as influencing the former) (Baumgaertner, Carlisle & Justwan (2018). Also, ‘complacency’ accounted for why some delayers and refusers were reluctant to be vaccinated. This complacency took the form of a perception of low personal risk and a valuing of “natural immunity” and might be better thought of as a form of ‘lay knowledge’ or ‘medical folk wisdom’ (Popay & Williams 1996; Motta & Calaghan 2020). As such, rather than constructing it as complacency per se, it might be more productive to focus on constructing and communicating vaccination as a collective act – one that even those at relatively low personal risk from serious COVID-19 illness should do to protect others

Similarly, the Vaccine Hesitancy Determinants Matric holds that contextual, individual and group, and vaccine-specific issues all impact the extent to which people are accepting of or hesitant towards vaccination (SAGE 2014). For instance, vaccination as a social norm was found to be an important individual and group influence. We found that a major barrier in the context of COVID-19 is the existence of conspiracy theories and Covid echo chambers.

Thus, reducing the circulation and belief in conspiracy theories will likely help control the spread of COVID-19, (Romer & Jamieson 2020) including in this case through potentially increasing vaccine uptake, perhaps particularly amongst BAME communities. Research suggests that people may be drawn to conspiracy theories when they promise to satisfy epistemic (e.g. desire for certainty), existential (e.g. a desire for control) and social (e.g. a desire to ‘fit in’ within a group) motives (Douglas et al 2017). BAME communities may be particularly at risk from a lack of knowledge and safety, because of their historical marginalization in society and because of the fact that morbidity and mortality from COVID-19 has been higher. Research suggests that experiences of ostracism, including due to an individual’s race or ethnicity, may lead to greater belief in conspiracy theories, perhaps as a defence mechanism (Douglas et al 2017; Crocker et al 1999). Further research on the role of ‘Covid echo chambers’ is needed. Our findings suggest that some people are reluctant to engage with others who hold, or may hold, differing opinions – particularly since COVID-19 policy is such a divisive and emotionally-charged issue. This lack of communication across echo chambers can have an ‘opinion reinforcing’ effect. (Garrett 2009). In the context of COVID-19 vaccination, strongly ‘pro-vaccination’ advertising or opinions may even be having a counter-productive effect for some, encouraging people to ‘double-down’ on their opposition or adding to their hesitancy. As such, working with individuals and communities, engaging with rather than criticising or dismissing their concerns (both legitimate and illegitimate) via mutually respectful dialogue is essential (Burgess et al 2021).

Vaccine delay may be usefully understood in relation to broader conceptual models on ‘patient delay’ (Walter et al 2012). In the context of COVID-19 vaccines, ‘appraisal delay’ (or decisional delay) (Andersen et al 1995) can be thought of as the question of ‘should I get vaccinated’? In order to reduce decisional delay, it is particularly important for policy and health organisations to address informational barriers, including some people’s perceived lack of information, the existence of conspiracy theories, and the existence of ‘Covid echo chambers’. For our participants, the lack of information was seen to be partly due to the fact that COVID-19 is a novel disease (i.e. scientists don’t yet have enough information) and due to the fact that insufficient or unclear information was being communicated to them (by e.g. medical or political agencies or individuals). Also, some felt that the information around vaccines and their efficacy or safety was at times too complex to understand, especially in light of new developments. This is perhaps another example of a phenomenon identified throughout the pandemic, referred to as ‘alert fatigue’. This is where frequently changing information (e.g. policies, guidelines, advisories) becomes increasingly difficult to interpret, comprehend and retain for members of the public (Williams et al 2020c). Also, utilization delay (as a form of behavioural delay) can be understood as the time between when a person decides they will need medical intervention (i.e. a vaccine) and the deciding act on that decision (i.e. to get vaccinated) (Safer et al 1979). Delayers in this sense had already decided they were going to “need” to be vaccinated. Utilization delay entails the individual asking themselves, is the medical care (i.e. vaccination) worth the costs? In this case, many delayers had decided that the benefits of vaccination – as a ‘passport’ to international travel were worth the perceived costs (e.g. the perceived infringement of their right to refuse a vaccine or concerns over potential future side effects). The decision to get vaccinated was perceived as not an entirely voluntary one, but one into which they were being ‘nudged’ or even indirectly forced - via the assumption that vaccine passports would be required in the future, especially for international travel.

Our findings reaffirm that vaccine passports are a controversial and divisive topic in the UK, in keeping with an apparent split in academic and political opinion (Drury 2021). Our participants also framed the issue as being one that is problematic in terms of access, equity and ethics (Osama, Razai & Majeed 2021; Hall & Studdert 2021). However, from a pragmatic public heath perspective, our findings suggest that, at least as far as international travel is concerned, they might increase vaccine uptake, or reduce vaccine delay, and that many assume they are a *fait accompli*. Thus, our data reflect recent public opinion surveys in the UK, that suggest a majority of the British public are supportive of vaccine passports, particularly for international travel (Ipsos Mori 2021b; Ibbetson 2021).

There is very little research on post-vaccine behaviour, including adherence to measures, in part due to the speed at which the vaccine has been rolled out in the UK. However, one theory, discussed in the academic and grey literature, as well as in government and the media, was that having a vaccine might lead to ‘complacency’, that is reduced adherence, regarding social distancing measures (Williams et al 2020c; The Lancet 2021; Cooper 2021) This was of public health concern, particularly if it were found to be the case that those having one dose of the vaccine were adhering less (since they themselves would not be fully protected and may also be at risk of transmitting the virus to others). However, our findings suggest this concern is likely not being borne out, and most people are likely adhering in much the same way as they did prior to getting vaccinated. This finding triangulates with recent survey evidence which suggests that a majority are not changing their adherence behaviour directly as a result of being vaccinated.

There are a number of limitations to note. Firstly, as with all qualitative studies, the generalizability of the findings is limited. As such, percentages in each vaccine group (accepters, delayers, refusers) should be viewed cautiously and only in association with larger quantitative surveys on the topic. Another limitation of the study is that although attempts were made to recruit and include as diverse a sample as possible, there is a relative underrepresentation of older adults (aged 50+ in the sample). However, for the purposes of the research question around vaccine uptake having a younger sample may be of benefit. Further strengths and limitations of the overall methodology and recruitment in the wider study are discussed in prior publications (Williams et al 2020a; Williams 2020b). However, a particular strength of this study is its ability to provide in-depth and nuanced context as to the reasons for vaccine acceptance, delay or refusal, as well as the facilitators and barriers to uptake and views on vaccine passports and post-vaccine adherence. COVID-19 policy is a rapidly changing landscape, and thus public attitudes to COVID-19 vaccines may evolve accordingly. Further research is needed to explore the evolution of attitudes to vaccines, as is comparative work comparing across various countries.

## Data Availability

Ethical restrictions related to participant confidentiality prohibit the authors from making the data set publicly available. During the consent process, participants were explicitly guaranteed that the data would only be seen my members of the study team. For any discussions about the data set please contact the corresponding author, Simon Williams.

